# Excess mortality analysis for Germany for all three COVID-19 waves in 2020 - 2021

**DOI:** 10.1101/2021.07.06.21260105

**Authors:** Alexej Weber

## Abstract

**Background and Aims:** The excess mortality has been used as a metric to estimate the impact of COVID-19 across countries. For Germany, we observe that during the second half of the first and second COVID-19 waves, the COVID-19 deaths are significantly higher than the excess mortality. We attribute the difference to the pre-dying effect. We then compare the excess mortality to the official COVID-19 death numbers and calculate the infection fatality rates (IFRs) and the percentage of infected individuals from excess mortality for different age bands. We also compare the impact of COVID-19 to past influenza waves and analyze the vaccination effect on excess mortality.

**Methods:** We forecast the baseline mortality from official data on deaths in Germany. Distributing a part of excess mortality into the near future, we lower the baseline simulating the pre-dying effect. From the observed mortality deficit, we estimate the percentage of infected individuals and then estimate the age-dependent IFRs.

**Results:** In the first wave, we find an overall excess mortality of ca. 8 000. For the second wave, the overall excess mortality adds up to ca. 56 000. We find, that the pre-dying effect explains the difference between the official COVID-19 deaths and excess mortality in the second half of the waves to a high degree. Attributing the whole excess mortality to COVID-19, we find that the IFRs are significantly higher in the second wave. In the third wave, the overall excess mortality is ca. 5 000. We find an excess mortality in mid-age bands which cannot be explained by the official COVID-19 deaths. For the senior band 80+, we find results in favor of a strong and positive vaccination effect for the third COVID-19 wave.

**Conclusions:** We conclude that in the first and second COVID-19 waves, the COVID-19 deaths explain almost all excess mortality when the pre-dying effect is taken into account. In the third wave in 2021, the excess mortality is not very pronounced for the 80+ age band, probably due to vaccination. The partially unvaccinated 40-80 age group experiences a pronounced excess mortality in the third wave while there are too few official COVID-19 deaths to explain the excess. The no-vaccination scenario for the 80+ age band results in a similarly high excess mortality as for the more younger age bands, suggesting a very positive vaccination effect on reduction of COVID-19 deaths.

## Introduction

To assess the impact of the COVID-19 pandemic, different metrics are used. Most prominent ones are the reported COVID-19 case and death numbers. Both metrics are problematic [1-3] as they strongly depend on the testing strategy (e.g. who and under which circumstances is tested), testing availability (e.g. expanding testing strategy) and the definition of COVID-19 deaths (PCR-confirmed or also only suspected).

During the COVID-19 pandemic, the testing for the corona virus has been widely expanded in Germany. While only a few hundred thousand tests were performed for influenza in 2018/2019, around 35 million COVID-19 tests have been performed in 2020, see [4-5]. The increasing number of tests in the first half of 2020 makes it difficult to reveal the true development of the COVID-19 cases in Germany during that period.

Due to these problems, we consider excess mortality as a more objective indicator to assess the impact of the pandemic on the population. Excess mortality is defined as the difference between the actual mortality in a period (say a specific calendar week) and the expected mortality for this period. Whenever the difference is negative, the negative excess mortality is called mortality deficit.

To reduce the spread of the COVID-19 disease, Germany, like many other countries, introduced so-called non-pharmaceutical interventions (NPIs), including most restrictive NPIs (mrNPIs) policies like mandatory stay-at-home and business closure orders. Since the excess mortality may theoretically be not only due to COVID-19 but also due to the potentially harmful side effects of NPIs [6-15], in this work we want to compare the official deaths due to COVID-19 with the excess mortality for different age groups. We want to analyze whether the COVID-19 deaths explain the excess mortality, or whether they are smaller or bigger.

If the excess mortality is much higher than the official COVID-19 death counts, it does however not necessarily mean that the difference is due to the real manifestation of the side effects of NPIs. The official COVID-19 death counts may be incomplete, or the difference in excess mortality may be due to additional circulating diseases. However, a substantial difference will require further investigation.

If the official COVID-19 death counts are much higher than the excess mortality, then this may be explained by the following factors: the official COVID-19 death counts may include deaths where COVID-19 was not the cause of death (deaths with the disease) or the crisis-free base line needs to be modified. It may be modified by external factors like NPIs which potentially lower the base line as less people die in accidents. It may also be modified as, after a period of mortality excess, the mortality will be lowered due to the pre-dying (harvesting) effect [2-3, 16]. Since the mortality in weeks before the second wave is not lower than the baseline mortality, we focus on the pre-dying effect (observable also after past influenza waves) in this work, which is the dominating effect modifying the baseline mortality.

Considering the pre-dying effect enables us to estimate the portion of infected people during the disease wave from the mortality deficit, which follows the mortality excess. Knowing the number of infected people and the excess deaths enables us to estimate the infection fatality rates (IFR) for each age group. We compare the IFR for each age group calculated with the pre-dying effect in the first wave (in spring 2020) and the second wave (autumn/winter 2020-2021) of COVID-19 in Germany.

We also compare the official COVID-19 death counts amongst each other. Here, we rescale them to the highest number of deaths in the most senior age group. We would expect the death counts to more or less overlap when rescaled. We will discuss the substantial differences in the Results section.

Finally, we want to analyze the vaccination effect on mortality numbers. Here, we want to know at which point in time it may be responsible for the reduction of death numbers.

## Data

We used the mortality data (updated on 23 November 2021) and the population structure data from the statistical bureau of Germany [17]. The official COVID-19 death counts (updated on 25 November 2021) are taken from the Robert Koch institute (RKI) [5]. We used weekly data, as daily mortality data are not available for different age bands. In addition, the weekly data contain less noise compared to daily data, and potential reporting delays, especially on weekends and public holidays, are smoothed out.

## Methods

### Forecast of the baseline

To calculate the excess mortality, the mortality baseline has to be subtracted from the actual death counts. The mortality baseline represents the background mortality (expected mortality) in absence of crises events like influenza or heat waves. Often, the expected deaths are calculated by taking the average of several past years of data [16-21]. It is important, however, to account for trends in data. The statistical bureau of Germany for example, in their weekly overview reports, calculates the baseline mortality by simply taking the average of 2016-2019 mortalities [17,21] without taking any trends into account. Since the total mortality numbers have been increasing on average since ca. 2005 for Germany, this simple method leads to an overestimated excess mortality.

More advanced methods apply sophisticated statistical models to past years of mortality data [22-24]. However, these more advanced approaches may lead to a biased forecast of the baseline if influenza and heat waves are not properly treated. Since the influenza waves mostly occur around the beginning of the year and heat waves in the middle of the year, they do not cancel out. The statistical model interprets these waves as a regular pattern. Hence, their estimate of the expected deaths for such periods is higher leading to a smaller excess mortality.

In addition, after the period of strong influenza waves, the mortality numbers experience a deficit due to the harvesting effect. Taking these data points into account results in the bias of setting the baseline too low.

In this work, we construct the expected deaths from clean parts of the mortality data without strong influenza waves or heat waves, adjusted to the current level of deaths, accounting for a crisis-free baseline. We also exclude long mortality deficit periods after strong influenza waves. We use a hybrid model consisting of the average of two models.

In the first model of the hybrid model, we fit a model having the trend and the first order Fourier components, for different age bands (AB):

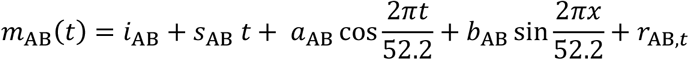

In this formula, *m* is the mortality depending on time *t* (discrete weeks). The parameters are: the intercept *i*, the slope *s* which accounts for the trend in the data, *a* and *b* are the coefficients of the first Fourier harmonics pair and *r* is the residual. The index AB stands for the age band: 80+, 60-80, 40-60 and 0-40.

We fit the model to the available weekly data of 2016-2021. We exclude the strong influenza and COVID-19 waves as well as outliers due to heat waves from the data. We also exclude the period of the strong mortality deficit after such waves. In the Results section, we provide more details on which data points have been taken to fit the model. To reduce the effect of negative auto correlations, we applied the moving average ma(5) model to the data prior to fitting the model. For the most noisy data of the 0-40 age band we apply ma(7).

In the second model of the hybrid model, the baseline is constructed as the average of the influenza-free years 2016 and 2019, adjusting them to the current mortality data [2,3] by rescaling. Here, the question arises of how to adjust the past data to the current level. One can either use the linear trend, as in the first part of the hybrid model, or to adjust the data using the age structure of the population (which can be found in [25]).

The disadvantage of using the available age structure data is that it is based on the census in 2011. From 2012 on, it is obtained from a forecast. This forecast uses different scenarios for migration, life expectancy and birth rate. If we adjust the mortality of a clean year 2016 to the level of the clean year 2019 using the age structure under basic scenario (scenario nr. 2 in the age structure data), we find good agreement for some age bands. We however find big discrepancies for some other age bands, although the adjusted mortality curves of mild years should coincide on average. Therefore, we do not use age structure data. Instead, to adjust the mortalities for the second model, we just take the number of deaths in 52 weeks in 2016 and 2019 and extrapolate this trend to 2020 and 2021. Here, instead of working with additive trends as in the first part, we work with rescaling to include multiplicative effects into the model.

Both methods depend strongly on the extrapolation of the trend to 2020 and 2021. Here, we consult the actual data in the beginning of 2020 and between both COVID-19 waves, and also the forecast from the age structure data, to decide whether we continue the trend seen in 2016-2019 to 2020, or not. Since we do not expect to have large jumps in age structure from year to year, this approach is the most reasonable one in absence of reliable age structure data. More details are given in the Results section.

The year 2020 started as a mild year, so we take the first weeks of mortality for 2020 before the first COVID-19 wave for the construction of the base year as well. Also, some influenza years are clean years if only younger age bands are considered. Therefore, we take more years of data into account for younger age bands.

We find that both parts of the hybrid forecast agree very well. In the above formula, we restrict ourselves to the first Fourier harmonics pair. The higher harmonics would fit smaller scale fluctuations. Here, the question arises where to set a cut-off. One can use either significance tests or information criteria. Different approaches lead to different cut-off numbers. Inclusion of higher harmonics will fit our model more and more to the average of corresponding data points in each week. Therefore, instead of applying higher order harmonics, we just average both models, which motivates the hybrid model. Thus, also additive and multiplicative effects are averaged.

The year 2020 started as a clean year, so the beginning of 2020 should correspond to the baseline. Therefore, we also adjust the baselines from each model to the first 10 data points of 2020 so that the baselines match the average mortality during the first 10 weeks. We include this adjustment into the error band around the baseline. We do not take the full adjustment however, but only 50%, since the beginning of 2020 may still start slightly below or slightly above the baseline.

To summarize, we use a hybrid model which is the average of two models. Model one: the first Fourier harmonics pair (fitted with a trend line to past mortality data (excluding influenza waves)) and model two: the average of clean mortality years, adjusted to the current level and smoothed out using a low order moving average.

### The pre-dying effect

Analyzing the weekly mortality data, we notice short term (period length of 1-10 weeks) and mid term negative autocorrelations (period length of 10-50 weeks). For example, the mortality excess during a short heat wave is followed by a mortality deficit after the heat wave. The broad mortality excess during a strong influenza wave in the beginning of the years 2017 and 2018 is followed by a long mortality deficit for the remaining year.

A natural interpretation is that during these excess mortality waves, a substantial part of excess deaths results from deaths from very weak individuals who would have died a few weeks later from other causes. Their deaths would form the baseline in the near future, but since they *pre-die* in the excess mortality wave, they fall out of the base line, which is then lowered for some period in the weeks following the mortality excess wave. The lowering occurs naturally also during the excess mortality period itself, leading to a greater mortality excess and greater number of people who die due to the disease or heat wave than actually seen when working with pure mortality excess.

From the lowering of the baseline, the percentage of infected individuals and the infection fatality rate (IFR) can be estimated, see [3].

As an example, let us assume that 10 000 elderly individuals die per week on average. During a heat wave week, suddenly 15 000 die. However, in the week following the heat wave, only 6 000 die. Then we can estimate that out of 5 000 excess deaths, 4 000 resulted from weak individuals with life expectancy of around 1 week. The remaining 1 000 deaths resulted from individuals with life expectancy longer than 1 week. The ratio 5 000/10 000 = 0.5 tells us that around 50% of the elderly population suffered from the heat wave (for the other 50% of individuals, maybe the weather was milder or they had better air conditions at home etc.), considering that heat is very deadly (fatality rate around 1) to very weak people. The percentage of 50% for the hit individuals is actually a lower bound and the fatality rate is the upper bound. It can also be that 100% of the elderly population suffer from the heat wave, and the fatality rate is 50%. In any case, we get good estimates of the fatality and the portion of the affected population.

To model the pre-dying effect for broader excess waves, we have to make two assumptions. One assumption is that the infection or heat wave causing the excess mortality is very deadly to very week individuals with short life expectancy. We assume a value of fatality close to 1. If the actual fatality is less, then the derived results for the number of infected and for the IFRs have to be rescaled like in the example above. The number of infected increases, the IFR decreases.

The second assumption is that the fatality depends strongly on the crisis-free remaining life expectancy of the individual - it will be smaller for individuals with longer life expectancy. Thus, we can model different fatality decays. In a previous work [3], we assumed a step function for the fatality: a constant fatality of around 1 was assumed for individuals with life expectancy of less than 20 weeks and a value much lower than 1 was estimated for individuals with longer life expectancy.

In this work, we want to model a more realistic decaying fatality. We assume an IFR of 1 for individuals with life expectancy of 1 week. So the individual dies during the first or second week of the infection (week *t*) instead of dying one week later (week *t+1*) from other causes. Then we introduce a decaying parameter *0<d<1*. The IFRs for individuals with life expectancy of *1, 2, 3, 4, …* weeks are then: *1, d, d*^*2*^, *d*^*3*^,…. We then distribute the excess mortality *EM* in a certain week *t* following this decaying pattern as

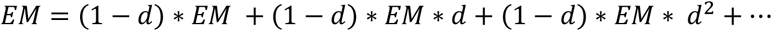

The first term then lowers the base line in week *t+1* by *EM*(1-d)*, in week *t+2* by *EM*d*(1-d)* and so on.

The algorithm to lower the base line then reads:

1. Start from week *n*_*0*_ where the excess mortality begins, lower the future base line using the formula given for a chosen parameter value 0.9 < d < 1.
2. Go to the next week and lower the lowered future baseline again using the excess mortality (with respect to the lowered baseline of the previous step)
3. Repeat step 2 as long as the mortality excess is existent
4. Fit the pre-dying baseline to the period with mortality deficit starting around 4 weeks after the end of the original mortality excess by varying the decaying parameter *d*.

The following modifications may improve the result: Instead of working with the original mortality curve, it is better to work with a smoothed curve using a lower order moving average, especially in the mortality deficit period. If we knew the exact number of deaths due to the disease, we might also fit the (pre-dying baseline + disease deaths) to the mortality curve. Since it is much harder to determine the number of people which died from the disease than the number of people which died in total, we did not include the COVID-19 death counts in the algorithm. However, not considering the COVID-19 deaths may lead to an underestimated number of people infected. In the apparent mortality deficit period, there can still be people dying from the disease. Not considering them will lead to an insufficiently lowered baseline when using the algorithm. To overcome this problem and at the same time, in order not to rely too much on official COVID-19 death counts, we also fit the (pre-dying baseline + official COVID-19 deaths) in the mortality deficit period of both waves. Although it is possible that a part of official COVID-19 deaths contains deaths “with” instead of from COVID-19, the number of reported COVID-19 deaths is much smaller in the mortality deficit period than in the middle of the disease wave.

We also fit the baseline to the mortality deficit period without taking the official COVID-19 deaths into account. This is being done to estimate the range of the number of infected and the range for IFRs.

It is also advisable to introduce a cut-off after about 30-50 weeks of baseline lowering, since the exact decay pattern may be different in the long term and additional effects may influence the excess mortality.

After the fitting the pre-dying baseline, the incremental portion of the infected in week *n* can be estimated from the excess mortality with respect to the lowered baseline as

(excess mortality with respect to the lowered baseline in week *n+2*)*(*1-d*)

divided by

(the value of the lowered baseline in week *n+1*).

In this formula, we assume a lag of 2 weeks from infection to death. Then the first factor corresponds to the number of individuals with life expectancy of 1 week dying from the disease in week *n+2*. The second term approximates the total number of weak individuals with life expectancy of around 1 week (here we can also use *n* instead of *n+1*, but the difference is negligible). For the younger age bands, the number of weak individuals are actually considerably smaller than the second factor, since the second factors also contains deaths from suicides and accidents which are proportionally bigger in the younger bands. So the calculated number of infected is in reality a lower bound and the calculated IFR the upper bound.

In this work, we define individuals with life expectancy of < 20 weeks as *weak*, and those with life expectancy of > 20 weeks as *healthy*. This is done in order to compare the results of this study with our previous study [3]. In [3], this was motivated from the observation that both the first COVID-19 disease period and the mortality deficit period lasted for about 10 weeks.

The total IFR for the age group can be estimated from the total excess mortality with respect to the pre-dying baseline and total number of infected individuals in the disease wave period. The IFR for weak individuals is assumed to be close to 1 following the decaying pattern *1, d, d*^*2*^ etc. depending on the remaining life expectancy. The IFR for healthy individuals can be estimated from the total mortality with respect to the pre-dying baseline multiplied by the factor of *d*^*20*^ (corresponding to the lowered base line in week 21, 22, …, see the formula for the sum of the tail of a geometric series) and the estimated total number of infected healthy individuals. The total excess mortality is then distributed with a factor of 1-*d*^*20*^ to weak individuals and with a factor of *d*^*20*^ to healthy individuals.

Of course, the number chosen (20) is a pure convention. We chose 20 because during the first wave the excess mortality lasted for around 10 weeks and was followed by a mortality deficit of around 10 weeks and the IFR and total number of infected can be estimated with even simpler considerations like in [3]. Our model is a very simple one and the precise decay of the IFR may be different in the short and long term. However, we aim to estimate the IFR on average and derive qualitative and quantitative average results for the disease period, so that the exact dependence of IFR on life expectance is not very important. We therefore also stop our calculation of the pre-dying base line after 30 weeks and calculate the pre-dying baseline for the first and second wave separately.

### Shape of the excess mortality and COVID-19 death numbers

During a disease wave, we expect the weak elderly individuals to start to die earlier and we also expect their excess mortality to end earlier than that of other age groups. The younger individuals are stronger on average, so they fight against the disease longer. We therefore analyze the official COVID-19 deaths to see whether they share this pattern. If the pattern is reversed, than this may be explained for example as follows. Either the younger age group was infected much earlier, so they die much earlier – or a substantial part of official COVID-19 deaths contains deaths *with* the disease, meaning individuals most likely died from other causes but were classified as COVID-19 deaths because they were positively tested for COVID-19. Then the official COVID-19 deaths will correlate better with the background infection rate which lags the death numbers by a lag of around 2 weeks (1 week to show the symptoms after infection and 1 week to die after the symptoms started).

### Vaccination effect

Vaccinations started first for the most senior age band 80+. We want to estimate the effectiveness of the vaccine *η*and then analyze how the vaccination may have influenced the excess mortality. In order to analyze the potential vaccination effect, we consider the following model:

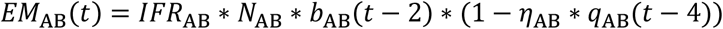

Here, for each age band (AB) we consider the excess mortality *EM*, at time *t* (in weeks) which is the product of the following terms. *IFR* is the infection fatality rate, *N* is the size of the population of the age band, *b* is the background newly infected ratio lagged with 2 weeks under the assumption of no vaccination, *η*is the vaccine effectiveness. Finally, *q* is the ratio of the vaccinated in the age band lagged with 4 weeks. The formula basically says that the excess mortality depends on the lagged number of infected times the IFR. If the vaccine has positive effectiveness to reduce the death numbers and infections, then the number of infected is smaller (with the lag of 2) and so the excess mortality gets reduced compared to the no vaccination case.

In order to estimate *η*from this formula, we have to know all other factors. The only true unknown is *b* which may be estimated from the unvaccinated younger age bands. But we do not need to estimate *b*. Dividing the excess mortality by the vaccination factor, we can estimate the hypothetic excess mortality *EM* which would be present had the age band been not vaccinated which is the same as assuming zero effectiveness:

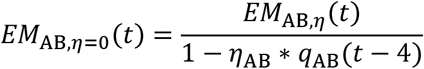

Setting the effectiveness *η*to 1, we can study the maximal possible vaccination effect, by applying this formula to the excess mortality of the most senior age band which was vaccinated first.

## Results

We perform the same analysis for different age bands and present the results in this section.

In Table 1, the results for the official COVID-19 death numbers, the excess mortality using the pre-dying effect, the estimate for the percentage of infected and the IFR for different age bands for the first and second disease wave are summarized. The numbers in the table are obtained as follows.

**Table 1.** Official COVID-19 death numbers, estimated death numbers from excess mortality applying the pre-dying effect, percentage of healthy individuals (life expectancy under normal conditions > 20 weeks) who died in the disease period, percentage of infected individuals and IFRs (total and for healthy individuals) for the first (calendar week 10-22 in 2020) and second COVID-19 waves (calendar week 41 in 2020 – 12 in 2021) in Germany. The confidence intervals are given in square brackets (calculated by shifting the baseline by 1 sigma up and down for the first wave and 2 sigma for the second wave). Larger shifts do not fit the mortality curve well in COVID-19-free times. The first line in each cell corresponds to the results taking the COVID-19 deaths into account in the mortality deficit period. The second line corresponds to not taking them into account. The values in bold correspond to the averaged values of the two methods. The values marked with * for the age band 0-40 and 40-60 are calculated by taking the nearest neighbor value from the next higher band since there was no pre-dying effect visible. For the age band “all” we calculate the numbers as for the other bands.

| Age band | Official COVID-19 Deaths (approx.) (in 1000) | Excess mortality (with pre-dying effect) (in 1000) | Deaths from healthy (%) | Infected during the wave (%) | IFR Total (%) | IFR healthy (%) |
| --- | --- | --- | --- | --- | --- | --- |
| 80+ (1<sup>st</sup> wave) | 5.6 | 7.7 (7.5,7.9)<br>6.8 (6.5,7.1)<br>**7.3 (7.0,7.5)** | 30 (20,40)<br>47 (35,59)<br>**38 (27,49)** | 4.1 (3.2,5.3)<br>2.4 (1.7,3.1)<br>**3.2 (2.5,4.1)** | 3.2 (2.4,4.2)<br>4.9 (3.6,6.9)<br>**3.8 (2.9,5.2)** | 0.98 (0.50,1.7)<br>2.4 (1.3,4.2)<br>**1.5 (0.8,2.6)** |
| 80+ (2<sup>nd</sup> wave) | 48 | 58 (57,60)<br>49 (48,50)<br>**54 (53,55)** | 47 (41,52)<br>68 (61, 74)<br>**56 (50,62)** | 20 (18,23)<br>8.7 (6.9,11)<br>**14 (12,16)** | 4.9 (4.2,5.7)<br>9.5 (7.6,12)<br>**6.5 (5.4,7.8)** | 2.4 (1.8,3.1)<br>6.7 (4.8,9.3)<br>**3.8 (2.8,5.0)** |
| 60-80 (1<sup>st</sup> wave) | 2.8 | 3.0 (2.9,3.3)<br>2.4 (2.4,2.5)<br>**2.7 (2.6,2.9)** | 8.8 (0,40)<br>22 (1.8,67)<br>**14 (0.55,52)** | 5.6 (2.1,15)<br>2.9 (0.8,7.5)<br>**4.1 (1.4,11)** | 0.29 (0.12,0.76)<br>0.46 (0.18, 1.7)<br>**0.36 (0.14,1.0)** | 0.026 (0,0.31)<br>0.10 (0.003, 1.2)<br>**0.051 (0.001,0.54)** |
| 60-80 (2<sup>nd</sup> wave) | 18 | 20 (19,22)<br>15 (13,16)<br>**17 (16,19)** | 38 (22,51)<br>76 (55,91)<br>**54 (35,68)** | 16 (12,22)<br>3.3 (1.2,6.4)<br>**8.7 (5.8,13)** | 0.70 (0.46,1.0)<br>2.5 (1.2,7.1)<br>**1.1 (0.7,1.8)** | 0.27 (0.10,0.52)<br>1.9 (0.64,6.5)<br>**0.6 (0.2,1.2)** |
| 40-60 (1<sup>st</sup> wave) | 0.4 | **0.34 (0.22,0.46)** | **14\* (0.55,52)** | **4.1 \* (1.4,11)** | **0.035 (0.01,0.14)** | **0.005 (0,0.073)** |
| 40-60 (2<sup>nd</sup> wave) | 2.1 | 2.4 (2.2,2.7)<br>1.6 (1.3,1.9)<br>**2.0 (1.8,2.3)** | 13 (1,33)<br>48 (13,77)<br>**26 (3.8,51)** | 16 (10,31)<br>4 (1.7,9.3)<br>**9.2 (5.4,19)** | 0.063 (0.03,0.11)<br>0.17 (0.062,0.47)<br>**0.093 (0.040,0.18)** | 0.0084 (0.0003,0.037)<br>0.082 (0.008,0.36)<br>**0.024 (0.0015,0.093)** |
| 0-40 (1<sup>st</sup> wave) | 0.04 | **0.006 (0.004,0.009)** | **14\* (0.55,52)** | **4.1 \* (1.4,11)** | **0.00041 (0,0.004)** | **0.000057 (0,0.00093)** |
| 0-40 (2<sup>nd</sup> wave) | 0.19 | **0.011 (0.002,0.063)** | **26\* (3.8,51)** | **9.2\* (5.4,19)** | **0.00033 (0,0.0033)** | **0.000087 (0,0.00012)** |
| all (1<sup>st</sup> wave) | 8.8 | 10.7 (10.5,11)<br>9.2 (8.7,9.7)<br>**9.9 (9.6,10.3)** | 24 (10,37)<br>43 (25,58)<br>**32 (16,47)** | 3.9 (2.8,5.9)<br>2.0 (1.4,3.1)<br>**2.9 (2.1,4.5)** | 0.33 (0.21,0.47)<br>0.55 (0.34,0.86)<br>**0.41 (0.26,0.60)** | 0.081 (0.017,0.13)<br>0.24 (0.086,0.51)<br>**0.13 (0.042,0.28)** |
| all (2<sup>nd</sup> wave) | 68 | 82 (79,83)<br>66 (63,67)<br>**74 (71,75)** | 44 (36, 49)<br>69 (60,75)<br>**56 (47,61)** | 17.8 (16,21)<br>6.2 (4.9,8.4)<br>**11 (9.8,14)** | 0.55 (0.44,0.63)<br>1.3 (0.90, 1.6)<br>**0.77 (0.60,0.92)** | 0.25 (0.16,0.31)<br>0.89 (0.55,1.2)<br>**0.43 (0.28,0.56)** |

In the mortality deficit period, we fitted the pre-dying base line to the actual mortality data both by using official COVID-19 deaths and without using them. If the official COVID-19 deaths are used in the mortality deficit period, then the baseline has to be lowered more – compared to the case where the COVID-19 death numbers are not used. If they are used, the calculated portion of infected increases and IFR decreases. We calculate both ways in order not to underestimate the IFR for each age band and in order to get a confidence interval. In addition, due to a higher total number of infected people after the disease wave, chances to detect someone dying with COVID-19 are much higher, and so is the risk for underestimating the IFR. The realistic values are given in bold as average between the two values (calculated by fitting the pre-dying base line to the actual mortality data both by using official COVID-19 deaths and without using them).

The total deaths in the pre-dying model are calculated as the total deaths in the excess mortality period with respect to the lowered baseline, so there are more total deaths than in the original excess mortality.

The aggregated excess mortality (using the pre-dying effect) for the first wave (week 10-22) is ca. 10 000 while we see around 9 000 COVID-19 deaths. This is due to the fact that the excess mortality started increasing a few weeks earlier than the official COVID-19 deaths. The original excess mortality (without the pre-dying effect) is ca. 8 000 (week 10-18) and ca. -1 500 (deficit, week 19-22). Thus, the net excess mortality without the pre-dying effect is ca. 6 500 for the first wave.

For the second wave, the excess mortality is ca. 75 000 while we have around 70 000 COVID-19 deaths. Therefore, either not all COVID-19 deaths were discovered, or people additionally died from other causes. The original excess mortality (without the pre-dying effect) is ca. 56 000 (week 41-7) and ca. -5 000 (deficit, week 8-12). Thus, the net excess mortality without the pre-dying effect is ca. 51 000 for the second wave.

In both waves, a large portion of deaths stems from 80+ individuals. The pre-dying effect explains the large difference between the original excess mortality and the official COVID-19 deaths by lowering the baseline. It also reveals that a large portion of COVID-19 deaths stem from weak individuals with short life expectancy of several weeks.

In the subsequent sections, we give additional figures to understand how the numbers in Table 1 were obtained. We also provided additional results and details.

### Model parameters

For each age band, we fit the linear trend model with the first pair of Fourier harmonics using clean past years of mortality data as described in the Methods section (first part of the hybrid model). The data is smoothed using the centered order 5 moving average to get rid of outliers and short term autocorrelations. For the age band 0-40 we increase the order of the moving average to 7, since the data is noisier. For the second part of the hybrid model we use the average of rescaled clean parts of past years. We provide the parameter values and standard deviations.

The raw data and the points used for fitting the first part of the hybrid model are shown in Figure 1. Both parts of the hybrid models were obtained as follows:

**Figure 1.**
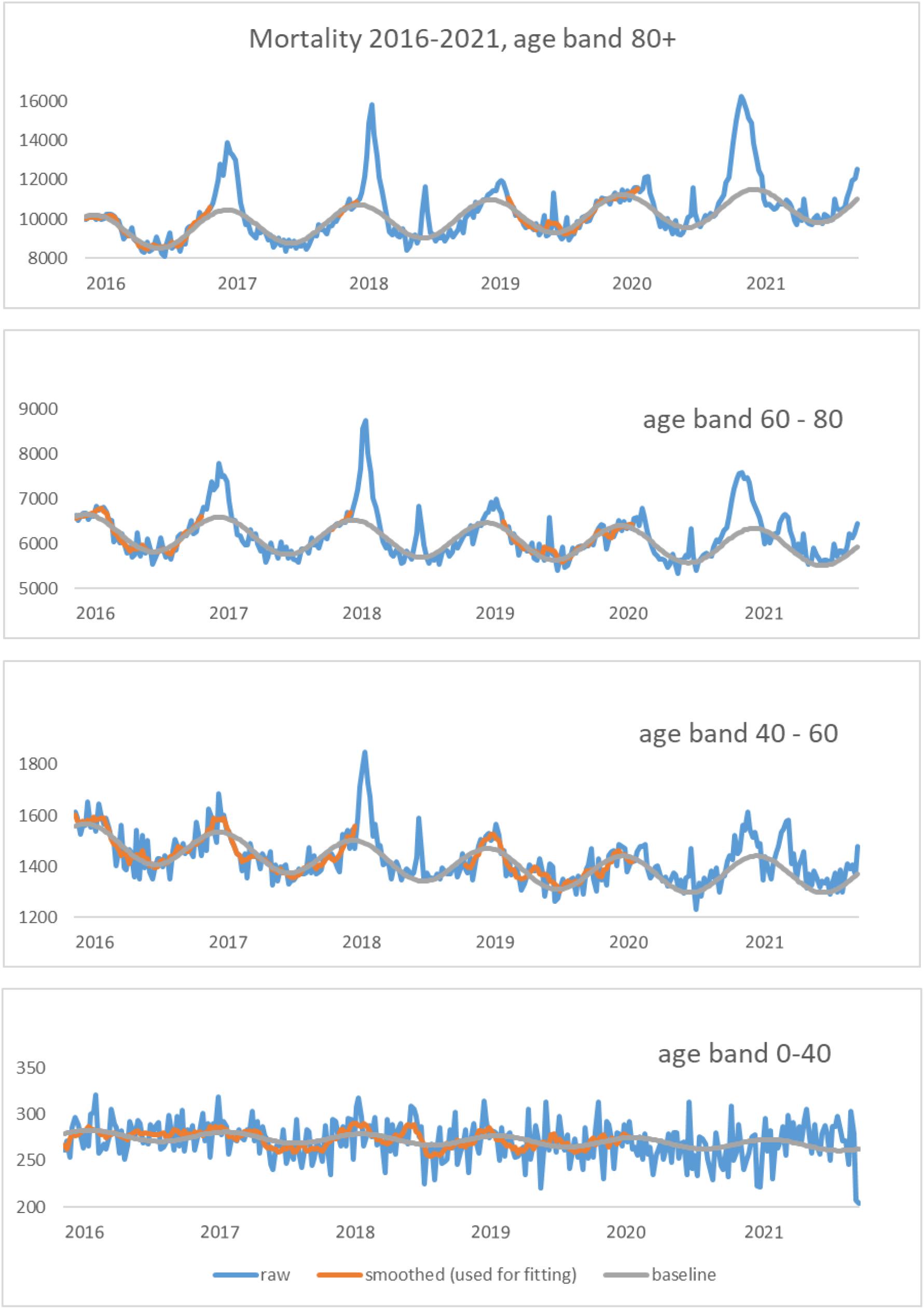
The figures show the mortality curves for different age bands of the population (blue). The smoothed points used to construct the baseline are shown in orange. The grey line represents the baseline of the first part of the hybrid model (trend and a pair of Fourier harmonics). We clearly observe average mortality deficits after strong mortality excesses for senior age bands, motivating the pre-dying effect.

#### Age band 80+

First part: intercept: 9271 (28), slope 5.0 (0.2), cosine amplitude 806 (24), sine amplitude 420 (25).

Second part: the average of the clean parts of the past years scaled with 2.9% (0.15%) per year (average yearly rate from 2016 to 2019). All parameters are highly significant.

The age structure data suggests a scaling of ca. 4% per year and leads to a too high baseline. Judging from the age structure forecast for 2020 and 2021, the positive trend should continue in 2020 and 2021 since this part of the population increases [25], so we keep the trend in 2020-2021.

#### Age band 60-80

First part: intercept: 6252 (16), slope -1.2 (0.1), cosine amplitude 332 (13), sine amplitude 228 (14).

Second part: the average of the clean parts of the past years scaled with -0.74% (0.1%) per year (average yearly rate from 2016 to 2019). All parameters are highly significant.

The age structure data suggests similar down scaling from 2016 to 2019, but then a stronger downscaling from 2020 onwards. We therefore continue the trend in 2020 and 2021.

#### Age band 40-60

First part: intercept: 1499 (4), slope -0.62 (0.033), cosine amplitude 57 (3), sine amplitude 45 (3).

Second part: the average of the clean parts of the past years scaled with -2.1% (0.13%) per year (average yearly rate from 2016 to 2019). All parameters are highly significant.

The age structure data suggests a less negative trend between 2016 and 2019. We set the slope to zero from 2020 (in order to match the mortality better between the COVID-19 waves). We took more points for fitting compared to more senior age bands as there is only one pronounced influenza wave in 2018. In this age band, we see a slight mortality excess during all of 2020. This could be due to NPIs taken by Germany or to a change in age structure of the population, such that this band gets bigger in size.

#### Age band 0-40

First part: intercept: 278 (1), slope -0.038 (0.008), cosine amplitude 1.32 (0.7), sine amplitude 5.4 (0.7).

Second part: the average of the clean parts of the past years scaled with -1.07% (0.24%) per year (average yearly rate from 2016 to 2019). All parameters (except for cosine amplitude, but it is small, so we keep it for consistency) are highly significant.

The age structure data suggests a positive trend between 2016 and 2021 (which however gets smaller), so we keep the negative slope in 2020 and 2021. We took all points from 2016-2019 for fitting, since there are no pronounced influenza waves in the data.

### Comparing COVID-19 waves to past influenza waves in 2017 and 2018

In this section, we compare the COVID-19 waves to past influenza waves. For simplicity reasons, we use only the first part of the hybrid model for the construction of the baseline. In the subsequent sections, we use the full hybrid model which leads to a slightly different baseline and consequently to slightly different excess mortality numbers for the COVID-19 waves. In Figure 1, we clearly observe that the mortality excess in 2017 and 2018 is followed by an average mortality deficit for some weeks, mostly pronounced in the more senior age bands, motivating the pre-dying effect.

Let us now compare the mortality excess with respect to the unadjusted baseline for 2020 and 2021 to the past influenza waves. For all age bands, the second COVID-19 wave was much bigger than the first one. We also see that the impact of COVID-19 on excess mortality correlates strongly with age. In the age band 0-40, no impact is directly visible. We also observe the mortality deficit after the second COVID-19 wave, which is interrupted by the third COVID-19 wave, however.

#### Age band 80+

The excess mortality in the second COVID-19 wave is bigger than for those of the past influenza waves, but it has the same order of magnitude. So in 2017, the excess mortality period corresponded to ca. 25 000 deaths. In the mortality deficit period ca. 9 000 died less. In 2018, the excess period counts ca. 25 000 deaths and the mortality deficit ca. 12 000 deaths. In the second COVID-19 wave, despite the national lockdown, the mortality excess counts ca. 43 000 deaths. In the first wave 2020 and in the third wave 2021, the mortality excess was ca. 6 000 and ca. 700, respectively.

#### Age band 60-80

The excess mortality in the second COVID-19 wave is bigger but comparable to those of the past influenza waves. So in 2017, the excess mortality period corresponded to ca. 8 000 deaths. In the mortality deficit period ca. 3 000 died less. In 2018, the excess period counts ca. 11 000 deaths and the mortality deficit ca. 4 500 deaths. In the second COVID-19 wave, the mortality excess counts ca. 13 000 deaths. In the first wave 2020 and in the third wave 2021, the mortality excess was ca. 2 000 and ca. 3 000, respectively.

#### Age band 40-60

The excess mortality in the second COVID-19 wave is comparable to those of the past influenza waves. So in 2017, the small excess mortality period corresponded to ca. 4 00 deaths. In the following mortality deficit period ca. 500 died less. In 2018, the excess period counts ca. 1 600 deaths with no substantial mortality deficit. In the second COVID-19 wave, the mortality excess counts ca. 1 200 deaths. In the first wave 2020 and in the third wave 2021, the mortality excess was ca. 200 and ca. 900, respectively.

#### Age band 0-40

Here, no substantial mortality excess is directly visible. When looking at smoothed numbers, a small influenza wave is visible in 2018. In 2021, we have a pronounced mortality deficit for this age band, probably due to NPIs taken by Germany. However, there is an ongoing mortality excess period in 2021 starting around cw 15-18. The total mortality excess for this period is ca. 400. The mortality excess period seems to end in cw 44. The last 2 data points for cw 44 and cw 45 are very likely to be updated however, so it is unclear whether this period excess has stopped in cw 44 in 2021.

We conclude that the mortality impact of the second COVID-19 wave for the 80+ age band was much higher than during the past two influenza waves, for the 60-80 the impact was comparable, and for the younger age bands the impact was smaller. In 2020 and 2021, the age band 0-40 experiences a mortality deficit on average (however, in cw 18 – 43 in 2021 there is a mortality excess).

### Comparing the COVID-19 deaths with excess mortality for the first COVID-19 wave

Now let us compare the official COVID-19 death numbers with the excess mortality in the first wave. For this, we add the COVID-19 death numbers to the baseline and also consider the pre-dying effect. The results are shown in Figure 2.

**Figure 2.**
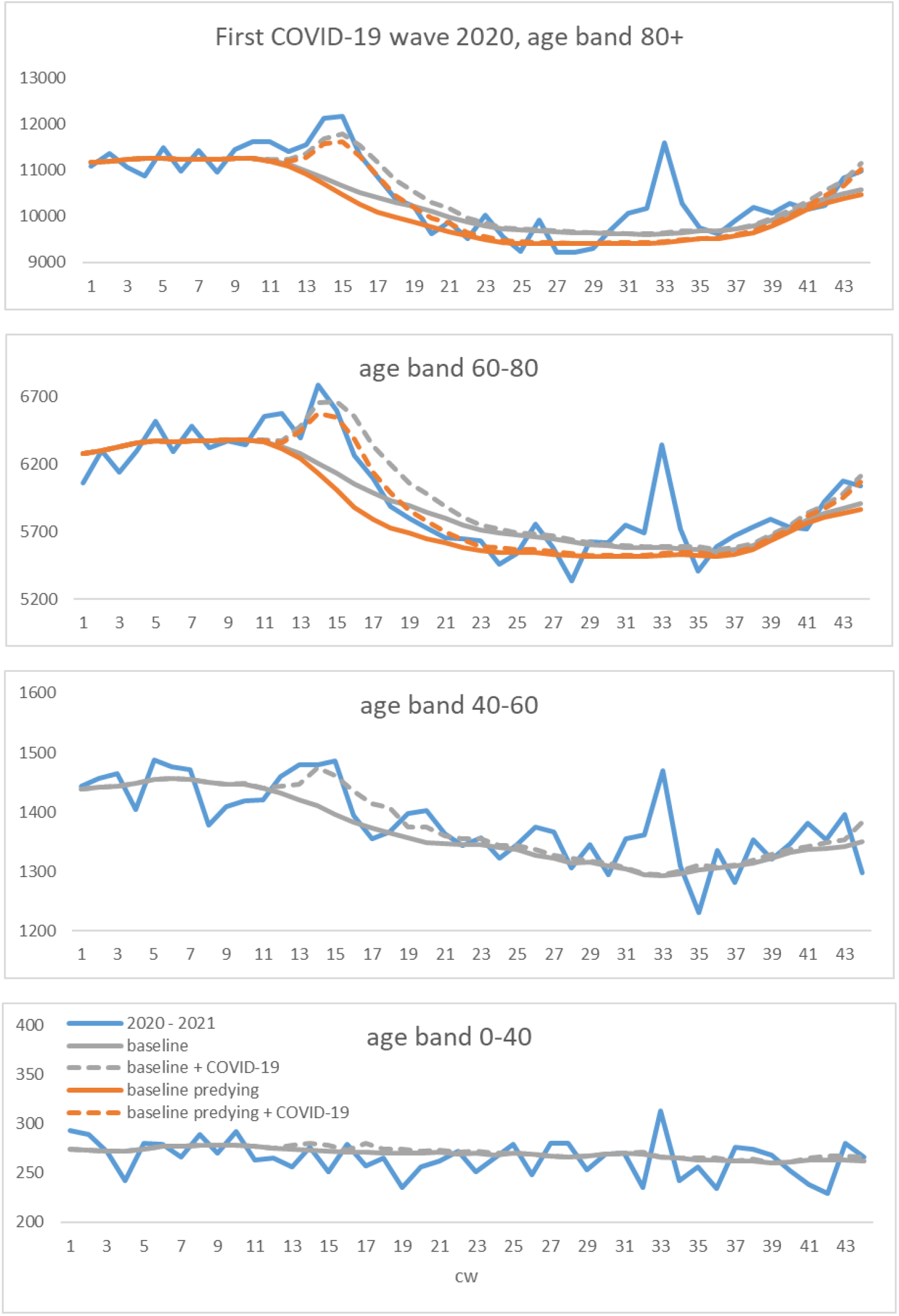
The figure shows the mortality curves for the different age bands (blue), the unadjusted baselines (grey), and the baselines + COVID-19 deaths (dashed grey). We see that the grey dashed line exceeds the mortality numbers in the second half of the wave. Applying the pre-dying effect (in orange), modifies the baseline so that COVID-19 deaths fit the mortality excess much better in the second half of the wave. Here we fit the decay parameter by fitting the baseline with COVID-19 death numbers to the mortality data in the deficit period. For age bands 0-60 we do not observe any mortality deficit, so we do not apply the pre-dying effect.

We see that the excess mortality started to appear first for the most senior band around calendar week (cw) 9 whereas the official COVID-19 deaths all start in cw 12. The reason can also be that another disease wave was circulating around cw 9-12 or in general the base line is slightly higher for this age band in that period. We have the same effect in the clean year 2016, where the mortality is higher than the base line in cw 9-12.

Only the age bands 80+ and 60-80 experience a mortality deficit after the disease wave, so we only apply the pre-dying effect there.

In cw 15 -25 the curve (baseline + COVID-19 deaths) is much higher than the actual mortality which experiences a mortality deficit from week 19 for the two most senior bands. If the baseline is lowered using the pre-dying effect to fit the mortality in cw 19-29, the pre-dying baseline + COVID-19 deaths explain the actual mortality curve much better in the excess period. Accounting the whole mortality excess to COVID-19 we can calculate the values in the corresponding rows in Table 1.

We see that for the 3 senior age bands, the official COVID-19 deaths do not explain the excess mortality in the first half of the wave fully. Probably the testing was expanded a few weeks too late. Actually the COVID-19 deaths explain the excess mortality better if the latter is smoothed using the low order moving average model. Therefore, either the mortality data or the COVID-19 death numbers probably contain some reporting mismatches.

Now let us calculate the mortality excess (around cw 9-19), the mortality deficit (around cw 20 -30), and the number of deaths in the heat wave in 2020 (around 31-35) for the two senior age bands using the unadjusted baseline. The result reads:

**Age band 80+**: mortality excess: ca. 6 000, mortality deficit: ca. 2 500, heat wave: ca. 4 000.

**Age band 60-80**: mortality excess: ca. 2 000, mortality deficit: ca. 1 000, heat wave: ca. 1 000.

We conclude that the net mortality excess was comparable to the number of additional deaths during the heat wave period. However, a larger number of people experienced the heat wave than were infected.

At this point, let us take the pre-dying effect into account and calculate the mortality excess (in cw 10-22). Now the mortality excess is bigger than official COVID-19 death numbers:

**Age band 80+**: mortality excess (using pre-dying effect): ca. 7 000, official COVID-19 death numbers: ca. 5 600.

**Age band 60-80**: mortality excess (using pre-dying effect): ca. 3 000, official COVID-19 death numbers: ca. 3 000.

We conclude that in the beginning of the wave, probably not all COVID-19 deaths were detected. Then in the second half of the wave, the COVID-19 deaths explain the excess mortality very well when the pre-dying effect is taken into account. Without the pre-dying effect, the COVID-19 deaths do not fit the much smaller excess mortality. The pre-dying effect also reveals that a large portion of COVID-19 deaths stem from weak individuals with short life expectancy of several weeks. The impact of the first COVID-19 wave on Germany was much smaller compared to other countries. The net number of deaths corresponded to the excess mortality during a regular heat wave.

Now let us provide the numbers for the 0-40 and 40-60 age bands:

**Age band 40-60**: mortality excess: ca. 340, official COVID-19 death numbers: ca. 370.

**Age band 0-40**: mortality excess: ca. 6, official COVID-19 death numbers: ca. 40.

### Comparing the COVID-19 deaths with excess mortality for the second COVID-19 wave

Now let us perform the same exercise we did in the previous section for the second COVID-19 wave, which hit Germany very strongly. In Figure 1, we observe that compared to the baseline, the extent of the excess mortality correlates with age. For more senior bands, the excess mortality is more pronounced.

We compare the official COVID-19 death numbers with the excess mortality in the second wave. For this, we again add the COVID-19 death numbers to the baseline and also consider the pre-dying effect. The result is shown in Figure 3.

**Figure 3.**
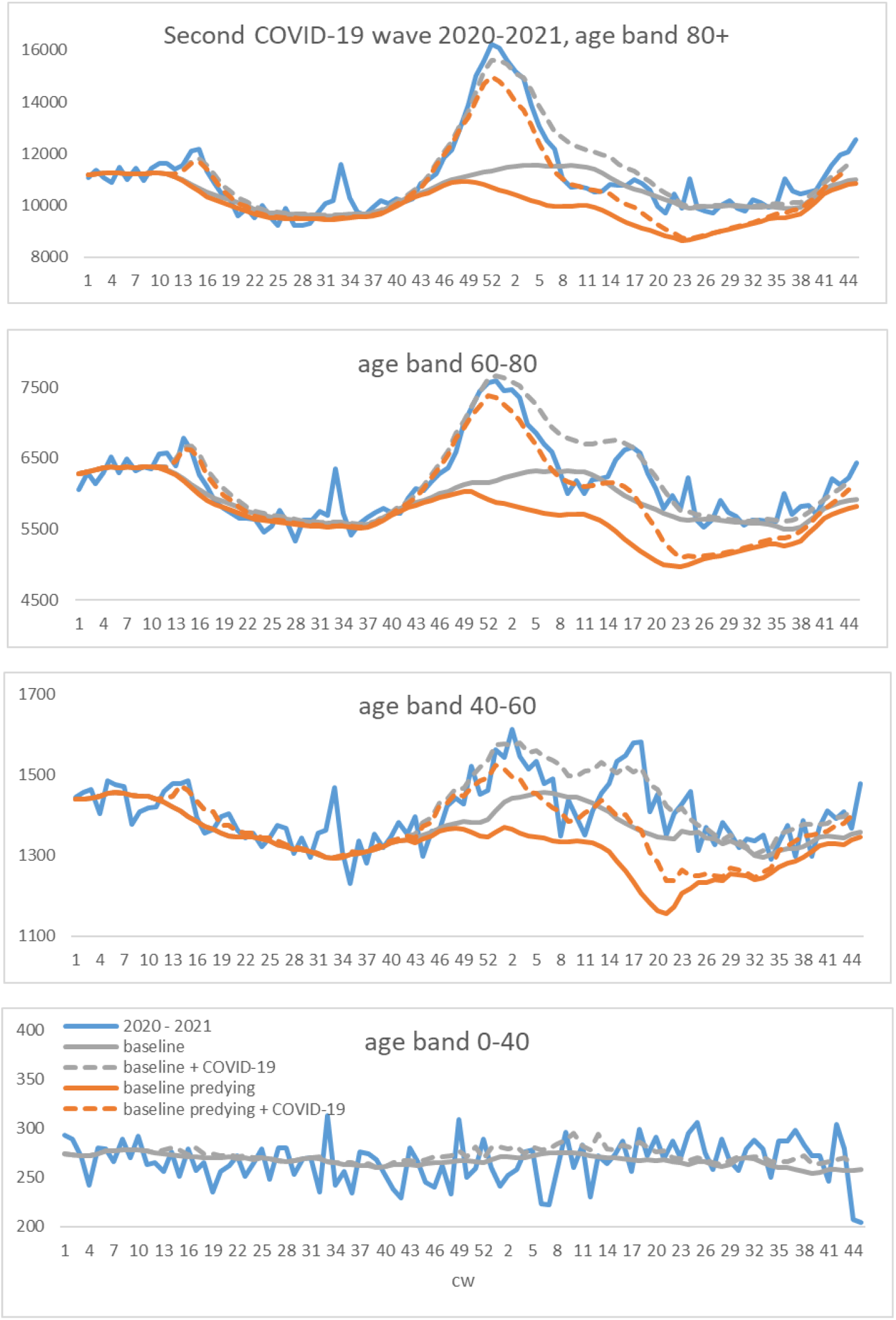
The figures show the mortality curves for the different age bands (blue), the unadjusted baselines (grey), and the baselines + COVID-19 deaths (dashed grey). We see that the grey dashed line exceeds the mortality numbers in the second half of the wave. Applying the pre-dying effect (in orange), modifies the baseline so that COVID-19 deaths fit the mortality excess much better in the second half of the wave. In this figure, we fit the decay parameter by fitting the baseline *with* COVID-19 death numbers to the mortality data in the deficit period. For the age band 0-40 we do not observe any mortality deficit, so we do not apply the pre-dying effect.

We see that the excess mortality started to appear for 2 senior age bands (but not the junior ones) around week 43 simultaneously. For the age band 40-60, it started ca. 2 weeks later. The COVID-19 deaths (all starting to increase around week 41) slightly exceed the excess mortality in the beginning. Here either our baseline has a small bias (is too high) or the COVID-19 deaths contain a small portion of regular deaths with COVID-19. Since this effect is observable for all age bands, we favor the second interpretation.

If the baseline is not adjusted with the pre-dying effect, the COVID-19 deaths explain well the first half of the excess mortality, but in the second half, they are significantly higher than the actual excess mortality.

Applying the pre-dying effect lowers the baseline, so that the COVID-19 deaths explain the second half of excess mortality much better. Again, we fit the curves in the mortality deficit period only.

If a substantial portion of COVID-19 deaths in the mortality deficit period contain regular deaths with COVID-19, then our adjustment with the pre-dying effect is too big. We get too high numbers of infected and too low IFRs. In fact, these numbers can be considered as upper bounds for the number of infected and lower bounds for the IFR. Therefore, we recalculate the pre-dying effect without taking the COVID-19 deaths into account, thus obtaining results shown in Figure 4. Not using COVID-19 deaths provides us with lower bounds for the number of infected and upper bounds for the IFR. The results are listed in Table 1.

**Figure 4.**
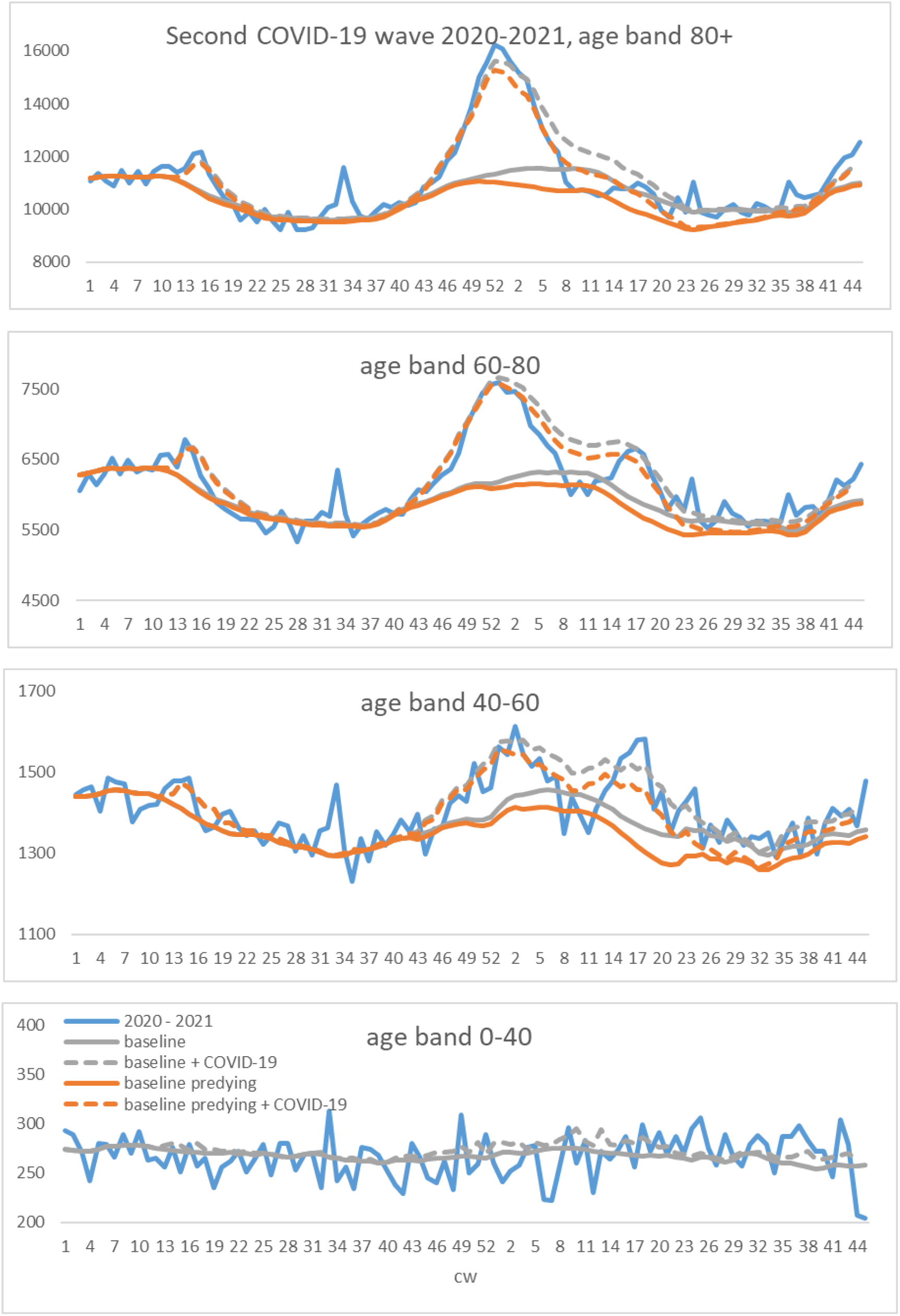
The figures show the mortality curves for the different age bands (blue), the unadjusted baselines (grey), and the baselines + COVID-19 deaths (dashed grey). We see that the grey dashed line exceeds the mortality numbers in the second half of the wave. Applying the pre-dying effect (in orange), modifies the baseline so that COVID-19 deaths fit the mortality excess much better in the second half of the wave. In this figure, we fit the decay parameter by fitting the baseline **without** COVID-19 death numbers to the mortality data in the deficit period. For the age band 0-40 we do not observe any mortality deficit, so we do not apply the pre-dying effect.

In Figure 3 and Figure 4 the pre-dying effect reveals that the excess mortality is substantially greater than the COVID-19 deaths. Here an explanation is that either not all COVID-19 deaths were discovered or reported, or the difference is due to other causes like e.g. the overload in hospitals.

In order not to underestimate the IFR, we account all excess mortality to COVID-19 however, calculating the values in Table 1.

Now let us compare the excess mortality with respect to the unadjusted baseline, the excess mortality using the pre-dying effect and the official COVID-19 death numbers in week 41 (2020) – 12 (2021):

**Age band 80+**: mortality excess (week 41 -7, unadjusted): ca. 42 000, mortality deficit (week 8-12):

ca. 4 000, excess mortality (with pre-dying): ca. 54 000, COVID-19 death numbers: ca. 48 000.

**Age band 60-80**: mortality excess (week 41 -7, unadjusted): ca. 13 000, mortality deficit (week 8-12):

ca. 1 000, excess mortality (with pre-dying): ca. 17 000, COVID-19 death numbers: ca. 18 000.

**Age band 40-60**: mortality excess (week 41 -7, unadjusted): ca. 1 200, mortality deficit (week 8-12):

ca. 200, excess mortality (with pre-dying): ca. 2 000, COVID-19 death numbers: ca. 2 000.

We conclude that the COVID-19 deaths explain the excess mortality very well when the pre-dying effect is taken into account. The pre-dying effect also reveals that a large portion of COVID-19 deaths stem from weak individuals with short life expectancy of several weeks.

Now let us provide the numbers for the 0-40 age band:

**Age band 0-40**: mortality excess: ca. 10, official COVID-19 death numbers: ca. 190.

### Comparing the excess mortalities for the third COVID-19 wave in spring 2021

In spring 2021, a third mortality excess wave starting around week 13 is visible for the pandemic in Germany, see Figure 1, Figure 3 and Figure 4. With the pre-dying baseline, the third wave is visible even better since the baseline is lowered. The wave gets proportionally bigger for younger age bands (with the exception of the most junior age band). This can be a vaccination effect, as the most senior age band got vaccinated first.

However, now there are not enough COVID-19 deaths to explain the excess mortality. Reasons can be undiscovered COVID-19 deaths, because less is tested or the COVID-19 deaths are still to be reported to RKI or other causes beside COVID-19.

### Comparing the COVID-19 death numbers and excess mortality across different age bands

Now let us compare the shape of the official COVID-19 death numbers and the excess mortality (with and without the pre-dying effect) for different age bands.

In case some numbers are still uncertain for the official COVID-19 death numbers, the RKI writes “<4” meaning a value between 0 and 4. We replace such entries with the value in between, namely 2.

In order to compare the numbers for different age bands, we rescale them to the age band for 80+ using the maximal number of deaths in the first wave. The result is shown in Figure 5. In this figure, we applied centered ma(2) smoothing to all age bands to reduce noise. This procedure also smooths out small potential reporting uncertainties and holiday effects.

**Figure 5.**
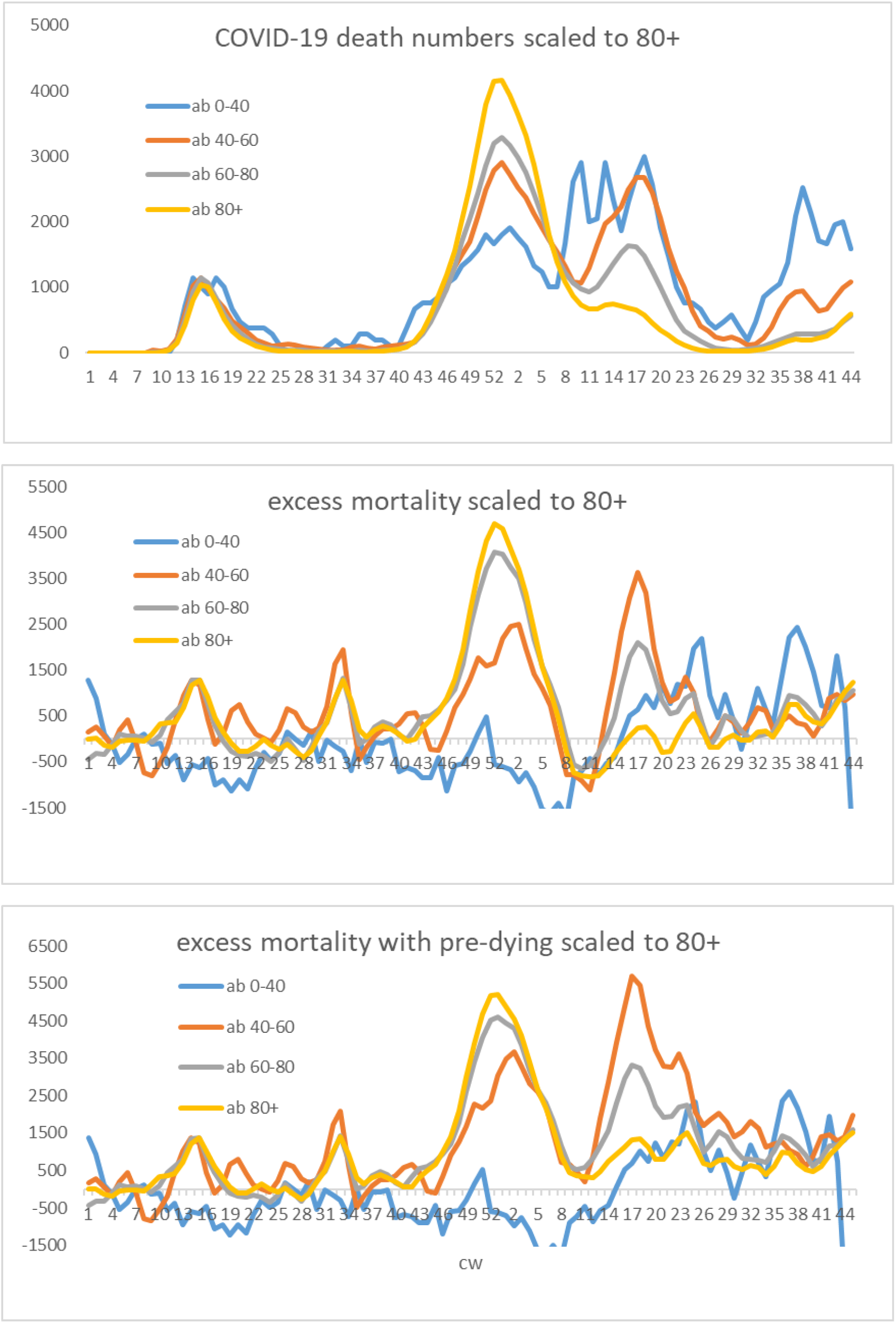
The figure shows official COVID-19 deaths, the excess mortality without the pre-dying effect and the excess mortality with the pre-dying effect, for different age bands rescaled to the 80+ age band in the middle of the first COVID-19 wave. We see that the curves share the same shape in the first wave. In the second wave the 80+ curve is highest. In the third wave, the COVID-19 deaths of the 80+ band are not very pronounced whereas the deaths for the 0-40 age band are proportionally highest. Yet, the third wave is better visible with the pre-dying effect. We see that proportionally the 40-60 age band is affected most in the third wave. The highest value of the excess mortality is even higher than in the second wave.

Concerning the excess mortality with respect to the pre-dying baseline: If we use the COVID-19 deaths in the fitting procedure, our adjusted baseline may get too low. If we do not use the COVID-19 deaths, the base line may be too high. In order to reduce the bias, we work with the average baseline using two methods, i.e.: to construct the pre-dying baseline, we calculated the average between the model taking the COVID-19 deaths into account and the model which didn’t, while fitting the pre-dying effect.

We see that the shapes look very similar except for the youngest age band 0-40 (here is no apparent correlation between the excess mortality and the COVID-19 deaths).

In the second COVID-19 wave, all other rescaled death numbers are smaller than the death numbers for the 80+ age band. This either means, that the second wave was more deadly to the senior population compared to the first wave, or that in the first wave, too few COVID-19 deaths from senior population were discovered.

For the second half of the first wave, in Figure 5 we observe that the COVID-19 death numbers for the 0-40 age band are higher than average. This could be due to individuals fighting longer against the disease. Then we observe that in the beginning of the second wave, the COVID-19 numbers for the 0-40 age band start increasing ca. 2 weeks earlier than average, and also start falling ca. 2 weeks earlier than average. This suggests that some of the deaths could be deaths “with” COVID-19 so that the numbers correlate with the background infection rate which lags the death numbers by ca. 2 weeks.

For the third wave, we again observe that the COVID-19 death numbers for the 0-40 age band start increasing ca. 2 weeks before the other bands on average start (the same is true for the fourth wave). We also observe a strong dependence of the numbers on age: all numbers are smallest for the 80+ band and biggest for the 0-40 age band. This can be the manifestation of the vaccination effect.

For the second half of the second wave, in the figure with the pre-dying effect, the excess mortalities for the 3 senior bands fall almost identically for all age bands, although the age band 80+ gets more vaccinated, suggesting that vaccination of 80+ was not the main cause for the reduction of excess mortality in that period.

For the second wave, we observe that the excess mortality for the 40-60 age band started to increase ca. 2 weeks later than the more senior bands, although the COVID-19 deaths start increasing in the same week for these age bands. This suggests that a part of the COVID-19 deaths for the 40-60 age band potentially contains deaths “with” COVID-19. Also, the highest point of the excess mortality for the 40-60 age band is ca. 2 weeks later as the ones for more senior bands. This could be due to factors like younger individuals fighting longer against the disease or hospitals working at their limits. For the COVID-19 deaths, all peaks occur in the same week.

The excess mortality of the 0-40 age band is unaffected by COVID-19. Shortly after the first wave it experiences a mortality deficit which stopped in week 25. In a similar way, we have a mortality deficit in the beginning of 2021 which stops around week 18.

Both age bands 40-60 and 60-80 experience a substantial mortality excess in the third wave where the highest value is ca. 50% less than for the second wave for the 60-80 age band, and where the highest value is bigger for the 40-60 age band in the third wave.

The third wave is better visible using the pre-dying effect. We note that the 40-60 age band is affected most, followed by the age band 60-80. The excess mortality for the most senior band is low probably due to the vaccination effect which we want to analyze in the next section.

### The vaccination effect on excess mortality

In this section, we want to analyze the largest possible effect of vaccination on the reduction of the excess mortality. We use the excess mortality with respect to the adjusted pre-dying baseline.

We want to analyze the vaccination effect on the reduction of excess mortality in the second wave and the increase of excess mortality for the third wave. In Germany, the age band 80+ was vaccinated first. Unfortunately, we do not have detailed data on the vaccination progress for different age bands. In the daily reports of the Robert Koch Institute from 14 March of 2021 [26], we find that in calendar week 11, around 52% of the age band 80+ have been vaccinated for the first time. The vaccination started around week 53 in 2020. Assuming a linear vaccination progress, we can estimate the function *q(t)* (see Methods section) of the vaccination ratio starting with 0% in week 53 in 2020 and being 52% in week 11. Assuming a vaccination effectiveness of *η*= 1, we can estimate the upper bounds for the excess mortality, had the age band 80+ not been vaccinated:

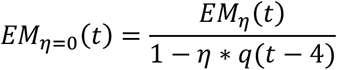

The result of the adjusted excess mortality for the age band 80+ is shown in Figure 6, together with other age bands. Under the assumption that the excess mortality was mainly caused by COVID-19 in weeks 1-11 in 2021, and assuming an effectiveness of 1 for the vaccination, the excess mortality of the age band 80+ would look like the dark blue line in Figure 6 without vaccination. We see that a few weeks after the start of vaccination, the modified excess mortality almost coincides with the original excess mortality because only a small portion of the age band had been vaccinated (*q<<1*). However, around week 12, assuming an effectiveness of 1, the vaccination effect becomes very strong. Without vaccination, the excess mortality would be proportionally similar to that of other age bands affected by the third wave.

**Figure 6.**
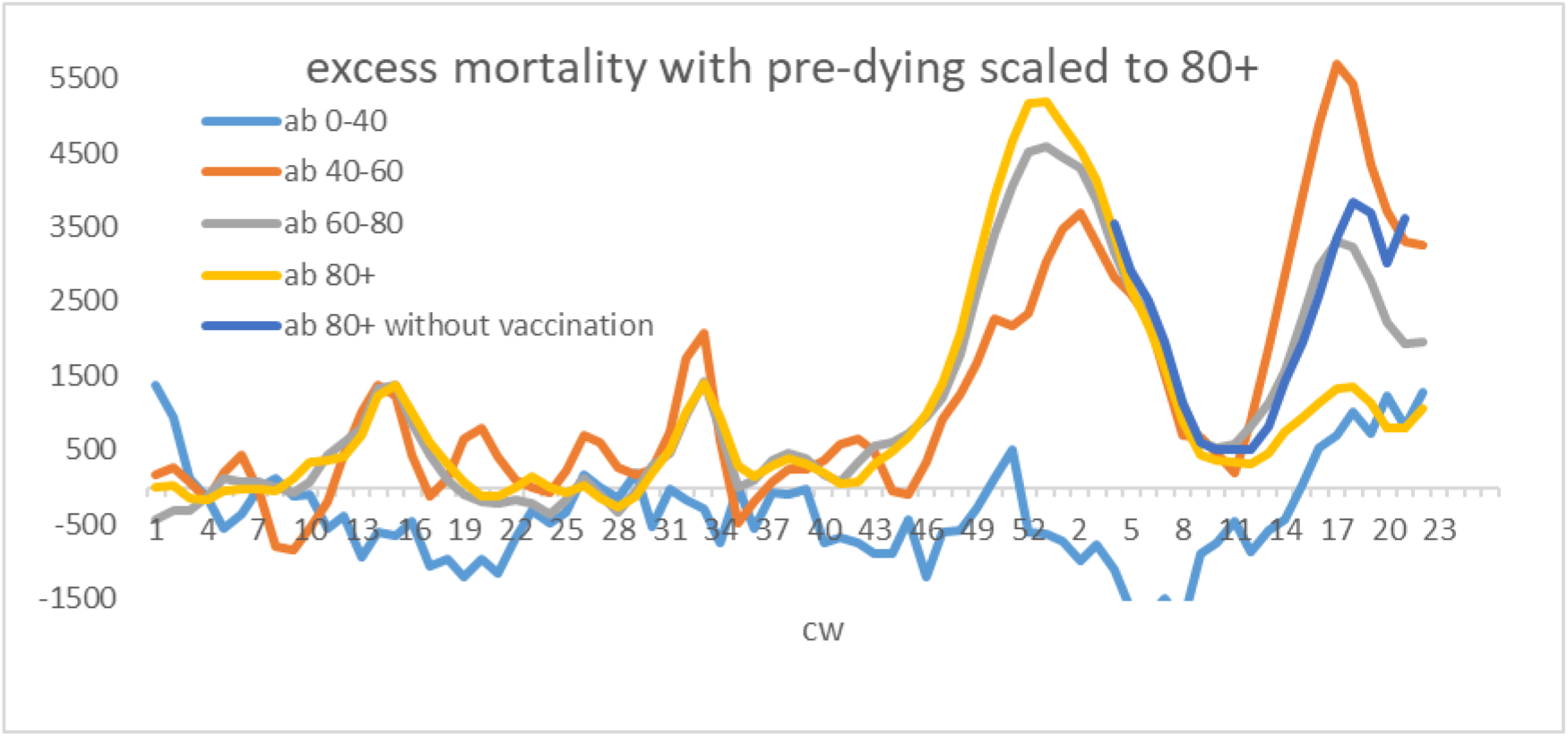
The figure shows scaled excess mortality curves for different age bands rescaled to the 80+ age band in the middle of the first COVID-19 wave. In addition, we have in dark blue the hypothetic scenario of the excess mortality for the unvaccinated 80+ age band assuming 100% effectiveness of the vaccine. We see that the excess mortality during the third wave for the age band 80+ gets significantly higher and is of the same order as for younger age bands. The vaccination effect on excess mortality in the second wave is very small, however.

To estimate the maximal number of lives potentially saved by vaccinations, we note that the excess mortality in week 4-12 in 2021 is ca. 12 000 using the pre-dying effect, see (Figure 6 yellow line). This number would increase by ca. 1 000 (assuming an effectiveness of 0.5) to 2 000 (assuming an effectiveness of 1) without vaccination (see dark blue line in Figure 6). Therefore, the vaccination would not have been the main cause of the reduction of excess mortality during the second wave, but it potentially prevented the age band 80+ from a stronger third wave and has an estimated effectiveness of 0.5-1 (if the excess mortality in the third wave is mainly caused by COVID-19).

As noted in previous sections, we still do not have enough COVID-19 deaths to explain the excess mortality in the third wave with respect to the pre-dying baseline. This may be due to a reporting delay and has to be observed carefully.

### Unexplained mortality excess

During our analysis, we noticed that the official COVID-19 death numbers probably start too late and do not capture the beginning of the first COVID-19 wave properly. We also noticed that during the second and third wave, not all excess mortality was explainable by COVID-19 death numbers.

We therefore subtract from the excess mortality calculated with the pre-dying effect (average of taking and not taking COVID-19 death numbers into account for fitting) the official COVID-19 death numbers to estimate the extent of this effect. Where the difference is large, the COVID-19 death numbers are either incomplete or the individuals died from other causes. The result is shown in Figure 7 for all age bands. The data has been smoothed with the centered ma(2) model.

**Figure 7.**
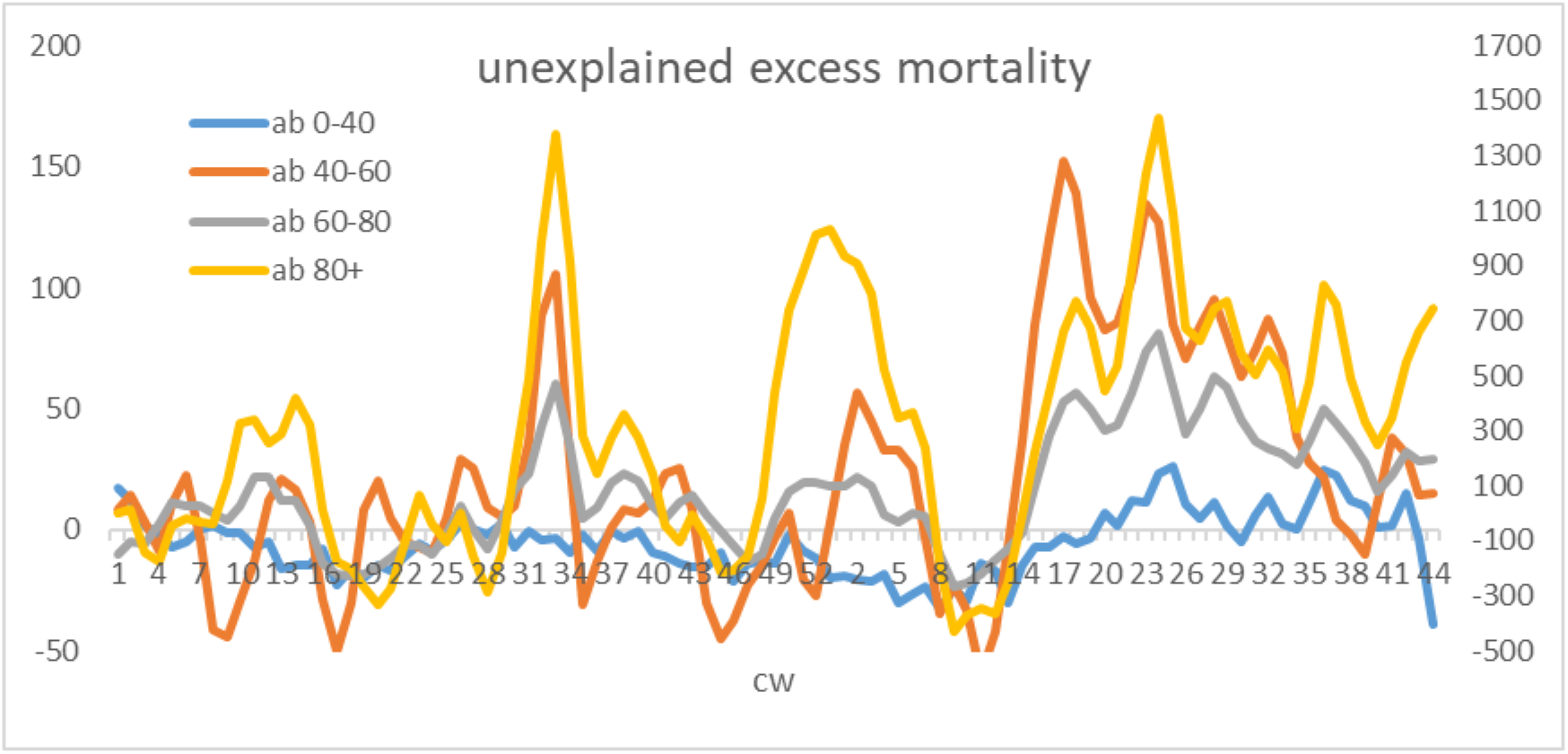
The figure shows unexplained mortality excess with the pre-dying effect (mortality excess minus COVID-19 deaths). One observes the mortality excess not explained by COVID-19 deaths in the beginning of the first COVID-19 wave (around week 13 in 2020), then at the heat wave (around week 32 in 2020), in the second COVID-19 wave (week 49 in 2020 – week 6 in 2021) and during the third wave. The age bands 0-40 (blue) and 40-60 (orange) have the axis on the left, the age bands 60-80 (grey) and 80+ (yellow) have the axis on the right.

Concentrating on values which are above the average noise level, we notice substantial excess mortality in several weeks of the years 2020-2021 not fully explained by COVID-19 death numbers:

First COVID-19 wave 2020: age band 80+ in week 10-14 and age band 60-80 weeks 11-13.

Heat wave 2020: age bands 40+ in weeks 31 – 34.

Second COVID-19 wave: Age band 60+ (week 49 - 7) and 40-60 (week 2 -6).

Third COVID-19 wave: age band 40+ starting from week 14 and 0-40 from week 20 (but the increasing started around week 14 as well).

The excess mortality during the heat wave does not have to be explained by COVID-19 numbers since we have similar outliers due to heat in past years in that period. The difference during the COVID-19 waves requires further investigation; the COVID-19 death numbers are probably incomplete.

## Conclusion

We observed that mortality excess is followed by a mortality deficit which motivates the pre-dying effect. In this work, we therefore studied excess mortality adjusting the baseline with the pre-dying effect. To construct the baseline in the first place, we applied a hybrid model taking several methods into account. This resulted in a more precise baseline than just adjusting the past mortality data with imprecise age structure data.

In the second halves of the first and second COVID-19 waves, we find a big discrepancy between the excess mortality with respect to the unadjusted baseline and the official COVID-19 death numbers. The COVID-19 death numbers are significantly larger than the net excess mortality. In this work, we explained the difference by the pre-dying effect. The net excess mortality in the first and second wave with respect to the unadjusted baseline is ca. 5 000 (ca. 8 000 excess, ca. 3 000 deficit) and ca. 52 000 (ca. 56 500 excess, ca. 4 500 deficit), respectively; with respect to the adjusted baseline (pre-dying effect), the mortality excess is ca. 10 000 and ca. 74 000, respectively; while the COVID-19 deaths amount to ca. 9 000 and ca. 68 000, respectively.

Taking the pre-dying effect into account, we were able to show that the official COVID-19 deaths explain almost all excess mortality. However, there are a few differences where we proposed the following explanation: There are probably too few official COVID-19 deaths in the beginning of the first wave; in the beginning of the excess mortality period (second wave) the official COVID-19 deaths exceed excess mortality most probably due to a substantial portion of deaths “with” instead of from COVID-19. However, as mortality excess increases, these deaths proportionally decrease and almost all excess mortality can be explained by the official COVID-19 deaths. During the second wave, the excess mortality is higher than the official COVID-19 death numbers however. This could be due to deaths from other causes and overload in hospitals or due to not detected COVID-19 deaths. In the end of the disease period (especially for the second wave), as in the beginning, a substantial portion of COVID-19 deaths are most probable due to deaths “with” COVID-19.

In both waves, a large portion of deaths stems from 80+ individuals. While the numbers almost agree for the adjusted excess mortality and official COVID-19 numbers, a noticeable portion remains unexplained, which could be either due to undetected COVID-19 deaths or deaths from other causes.

We analyzed past influenza waves and compared them to the COVID-19 waves. We found that COVID-19 in the second wave hit Germany stronger than past strong influenza waves.

Using the pre-dying effect, we calculated the number of infected during the disease period and the IFR for the two COVID-19 waves. We find that IFRs are higher in the second wave, which could for example be due to a mutated virus or to the overload in hospitals.

The obtained upper bound for the overall IFR (in %) for the first COVID-19 wave, 0.41 (0.26,0.60), agrees with IFRs found in the meta study [27] (0.23-0.27) and two analyses for Germany [28-29] (0.26-0.41) using seroprevalence data. It also agrees with our previous result (0.3-0.5) using a different decaying pattern for the pre-dying effect [3].

For the second wave, the upper bound for the overall IFR (in %) is estimated to be around twice as high, 0.77 (0.60,0.92), compared to the first wave.

For the first and second wave, the lower bound of the overall percentage of infected during the wave is estimated to be 2.9 (2.1,4.5) and 11 (9.8,14), respectively.

The official COVID-19 deaths for the age band 0-40 are proportionally smallest compared to other age bands, and they lag the deaths of other bands by a lag of 2, most probably due to a portion of deaths with COVID-19.

We find a mortality deficit in 2020 and beginning of 2021 for the 0-40 age band and a slightly increased mortality for the age band 40-60 in 2020 between the waves. This could be due to the changing age structure of the population or NPIs.

We notice a big mortality excess during and after the third COVID-19 wave which is much bigger than the official COVID-19 deaths for all age bands. This may be due to the reporting delays, but has to be further investigated.

We analyzed the vaccination effect for the 80+ age band. Vaccination most probably saved lives for a few thousands of individuals in the end of the second wave, but was not the main driver for the reduction of death numbers in that period. However, without vaccination, the death counts in the third wave would probably be proportionally as high as for the other bands, which suggests a very strong and positive vaccination effect with an estimated effectiveness of 0.5-1 for the 80+ age band.

## Data Availability

all data is publically available under the following links listed below

https://www.destatis.de/DE/Themen/Gesellschaft-Umwelt/Bevoelkerung/Sterbefaelle-Lebenserwartung/Tabellen/sonderauswertung-sterbefaelle.xlsx?__blob=publicationFile

https://www.rki.de/DE/Content/InfAZ/N/Neuartiges_Coronavirus/Projekte_RKI/COVID-19_Todesfaelle.xlsx?__blob=publicationFile

## References

1. Riffe T, Acosta E. Data resource profile: COVerAGE-DB: a global demographic database of COVID19 cases and deaths. International Journal of Epidemiology, 50(2):390–390f, 2021.

2. Weber A, Assessing the lockdown effect from excess mortalities. doi: 10.1101/2021.01.22.21250312

3. Weber A, Inferring the Prevalence and Age Dependence of Infection Fatality Rate from Excess Mortalities for COVID-19 in Germany (February 15, 2021). Available at SSRN: https://ssrn.com/abstract=3785836 or http://dx.doi.org/10.2139/ssrn.3785836

4. Bericht zur Epidemiologie der Influenza in Deutschland Saison 2018/19. Robert Koch Institut. https://edoc.rki.de/handle/176904/6253

5. Covid-19 Test numbers and test capacities for Germany. https://www.rki.de/DE/Content/InfAZ/N/Neuartiges_Coronavirus/Daten/Testzahlen-gesamt.html Official COVID-19 death counts. Robert Koch institute. https://www.rki.de/DE/Content/InfAZ/N/Neuartiges_Coronavirus/Projekte_RKI/COVID-19_Todesfaelle.html

6. Issue brief: Reports of increases in opioid related overdose and other concerns during the COVID pandemic. Accessed August 26, 2020. https://www.ama-assn.org/system/files/2020-08/issue-brief-increases-in-opioid-related-overdose.pdf

7. Guha-Sapir D, Moitinho de Almeida M, Keita M, Greenough G, Bendavid E. COVID-19 policies: Remember measles. Sills J, ed. Science. 2020;369(6501):261–261. doi:10.1126/science.abc8637

8. O’Leary ST, Trefren L, Roth H, Moss A, Severson R, Kempe A. Number of Childhood and Adolescent Vaccinations Administered Before and After the COVID-19 Outbreak in Colorado. JAMA Pediatrics. Published online December 7, 2020. doi:10.1001/jamapediatrics.2020.4733

9. Report 19 - The Potential Impact of the COVID-19 Epidemic on HIV, TB and Malaria in Lowand Middle-Income Countries. Imperial College London. Accessed August 26, 2020. http://www.imperial.ac.uk/medicine/departments/school-public-health/infectious-diseaseepidemiology/mrc-global-infectious-disease-analysis/covid-19/report-19-hiv-tb-malaria/

10. Kaufman HW, Chen Z, Niles J, Fesko Y. Changes in the Number of US Patients With Newly Identified Cancer Before and During the Coronavirus Disease 2019 (COVID-19) Pandemic. JAMA Netw Open. 2020;3(8):e2017267–e2017267. doi:10.1001/jamanetworkopen.2020.17267

11. Fragala MS, Kaufman HW, Meigs JB, Niles JK, McPhaul MJ. Consequences of the COVID-19 Pandemic: Reduced Hemoglobin A1c Diabetes Monitoring. Population Health Management. Published online June 29, 2020. doi:10.1089/pop.2020.0134

12. Wenham C, Smith J, Davies SE, et al. Women are most affected by pandemics — lessons from past outbreaks. Nature. 2020;583(7815):194–198. doi:10.1038/d41586-020-02006-z

13. Loades ME, Chatburn E, Higson-Sweeney N, et al. Rapid Systematic Review: The Impact of Social Isolation and Loneliness on the Mental Health of Children and Adolescents in the Context of COVID-19. J Am Acad Child Adolesc Psychiatry. 2020;59(11):1218-1239.e3. doi:10.1016/j.jaac.2020.05.009

14. Sher L. The impact of the COVID-19 pandemic on suicide rates. QJM. 2020;113(10):707–712. doi:10.1093/qjmed/hcaa202

15. Roelfs DJ, Shor E, Davidson KW, Schwartz JE. Losing life and livelihood: A systematic review and meta-analysis of unemployment and all-cause mortality. Social Science & Medicine. 2011;72(6):840–854. doi:10.1016/j.socscimed.2011.01.005

16. Alicandro G, Remuzzi G, La Vecchia C. Italy’s first wave of the COVID-19 pandemic has ended: no excess mortality in May, 2020. The Lancet. 2020;396(10253): E27–E28 doi:10.1016/S0140-6736(20)31865-1

17. Statistisches Bundesamt, https://www.destatis.de/

18. Krieger N, Chen JT. Excess mortality in men and women in Massachusetts during the COVID19 pandemic. The Lancet. 2020; 395(10240): 1829. doi: 10.1016/S0140-6736(20)31234-4

19. Modig K, Ahlbom A. EXCESS MORTALITY FROM COVID-19. WEEKLY EXCESS DEATH RATES BY AGE AND SEX FOR SWEDEN AND ITS MOST AFFECTED REGION. European Journal of Public Health, 2020;ckaa218, doi:10.1093/eurpub/ckaa218.

21. Zur Nieden F, Sommer B, Lüken S. Sonderauswertung der Sterbefallzahlen 2020. WISTA – Wirtschaft und Statistik, ISSN 1619-2907, Statistisches Bundesamt (Destatis), Wiesbaden, Vol. 72, Iss. 4, pp. 38–50. 2020.

22. European monitoring of excess mortality for public health action (EuroMOMO). Work package 7 report. A European algorithm for a common monitoring of mortality across Europe. Copenhagen: EuroMOMO; (Aaccessed: 20 Aug 2020).

23. Locatelli I, Rousson V. A First Analysis of Excess Mortality in Switzerland in 2020. medRxiv 2021.03.14.21253551; doi: https://doi.org/10.1101/2021.03.14.21253551

24. Karlinsky A, Kobak D The World Mortality Dataset: Tracking excess mortality across countries during the COVID-19 pandemic. medRxiv 2021.01.27.21250604; doi: https://doi.org/10.1101/2021.01.27.21250604

25. Statistisches Bundesamt, age structure. https://service.destatis.de/bevoelkerungspyramide/#!y=2021

26. RKI Tagesbericht 17 March 2021 https://www.rki.de/DE/Content/InfAZ/N/Neuartiges_Coronavirus/Situationsberichte/Maerz_2021/2021-03-17-de.pdf?blob=publicationFile

27. Ioannidis JPA, Infection fatality rate of COVID-19 inferred from seroprevalence data. Bulletin of the World Health Organization 2021;99:19–33F. doi: http://dx.doi.org/10.2471/BLT.20.265892

28. Streeck H, Schulte B, Kümmerer BM, Richter E, Höller T, Fuhrmann C, et al. Infection fatality rate of SARS-CoV-2 infection in a German community with a superspreading event [preprint]. Cold Spring Harbor: medRxiv; 2020. https://doi.org/10.1101/2020.05.04.20090076

29. Kraehling V, Kern M, Halwe S, Mueller H, Rohde C, Savini M, et al. Epidemiological study to detect active SARS-CoV-2 infections and seropositive persons in a selected cohort of employees in the Frankfurt am Main metropolitan area [preprint]. Cold Spring Harbor: medRxiv; 2020. https://doi.org/10.1101/2020.05.20.20107730

